# Mapping the Landscape of Community Care for Children, Young People and Families: A Co-Produced Project

**DOI:** 10.64898/2026.09.10.26362741

**Authors:** Sorcha Ní Chobhthaigh, Camille Cox, Twyla Greenway-Bailey, Josephine Musanu, Denaneer Khan, Amie Buhari, Mel Green, Cyra Neave, Mina Mawi, Rochelle A. Burgess, Delan Devakumar

**Affiliations:** Institute for Global Health, UCL, UK; Newham Community Project, UK; The Hebe Foundation, UK; The Open University, UK; The Anna Freud Centre, UK; London Borough School, UK

**Author notes:** Corresponding author: Sorcha Ní Chobhthaigh, Institute for Global Health, UCL, UK < >. These authors contributed equally as senior authors (Rochelle A. Burgess, Delan Devakumar).

**Keywords:** Community care, children and young people, mental health, service mapping, health equity, third sector, participatory research

## Abstract

In increasingly strained socio-political-economic contexts, reliance on community supports deepens, particularly for those failed by or unable to access appropriate care through statutory services. However, little is known about the scope, equity-oriented provision or the conditions that enable and constrain community care.

This mapping addressed these gaps, exploring community services in London and Greater Manchester, supporting children’s mental health and wellbeing. Co-produced with co-researchers and community stakeholders from conceptualisation to dissemination, we conducted a systematic scoping of community-based services, not government-provided or commissioned, offering free mental health or wellbeing support for children (0–18 years), carers, or families. We extracted service characteristics and equity-oriented provision, and provider and stakeholder insights informed a collaborative narrative synthesis.

We identified 297 eligible services (261 London, 41 Greater Manchester), with 48.9% of initially identified services excluded due to government commissioning/contracts. The majority (84%) advertised at least one equity-oriented practice, most commonly cultural safety/specific care (38%), though few explicitly intersectional (11%). Among those with a primary specialisation, support for violence and abuse was highest (12%). Services predominantly supported older children, with gaps in early intervention and prevention. The synthesis revealed trade-offs and tensions: responsiveness competes with planning, capacity-building and communications; navigation barriers alongside selective information sharing; and the ripple effects of funding precarity undermining provision and choice.

Community supports are relied on yet structurally vulnerable. As policy increasingly emphasises community partnerships, this requires sustainable funding models, collaboration guidance and resourcing to embed equity-oriented practices.

**Highlights:**

- Young people and community stakeholders co-led analysis and interpretation
- 84% of services advertised equity-oriented provision but few intersectional
- Prioritising responsive delivery forces trade-offs against planning and promotion
- Strategic invisibility is used to protect communities and prevent misunderstanding
- Funding precarity shapes provision, access and continuity of care

## Introduction

### What is community care?

Community care predates formalised state-funded systems and whether through community-based third sector services, community-led initiatives or ‘informal’ networks of care, continues to emerge in response to community need and local socio-political-temporal events. ‘*Community*’ in itself represents a spectrum of meaning, reflecting shared experiences, understandings or beliefs, collective goals or actions, geographic space or place(1). It can be anchored to a physical place and simultaneously transcend space, as a place of meaning, source of belonging or sense of mattering(2). Physical spaces within and for communities can offer places of informal care, shared environments that foster connection, and protected spaces for specific marginalised identities. They may also offer targeted supports, including pro-actively inclusive approaches or care for those who do not feel safe or cannot freely access state services. Unbounded by the parameters of clinical treatment, offerings may include youth work, alternative or holistic approaches, allowing for diverse conceptualisations of mental health and wellbeing, often centring connection, empowerment, and crucially, choice.

Where care is located, how it is delivered and who it is delivered by shapes who reaches it and how it is experienced. In mental health treatment, the quality of the therapeutic relationship and ‘match’ between those seeking support and providers is considered predictive of effectiveness(3), with feelings of safety, autonomy and choice - intertwined with feeling seen and understood - central to young people’s experiences(4,5). Mis-matches in understanding or approach may contribute to delays and barriers accessing appropriate care, less measurable improvement and broken trust(5,6). Community-embedded services attuned to the needs of their communities may hold expertise missing in state-provided care, understanding the socio-political-cultural realities faced by their members, being better positioned in place and power to build trust and navigate barriers to engagement. In turn, these services may be able to act as a trusted bridge to statutory services, facilitating service navigation, fostering system literacy and engagement if or when needed.

### The need for community care

Less than 10% of the National Health Service (NHS) budget is allocated to mental health, despite accounting for more than 20% of the disease burden (7), resulting in a significant gap and risk of escalating unmet need to crisis point. While recent health system policy orients more towards prevention (8) formalised services and resources have in practice prioritised treatment over early intervention. Once referred, high thresholds and lengthy waiting periods for accessing Child and Adolescent Mental Health Services (CAMHS)(9,10) amplify the gap between provision and need. These universal bottlenecks alongside barriers to help-seeking such as mental health literacy, misunderstanding and fear of judgement or repercussion(11) are compounded by inequities in referral and obtaining appropriate supports for racially and ethnically minoritised families(12–15). Even if these families access services, they continue to experience barriers at the point of treatment, including insufficient language translation, barriers associated with migration status, limited treatment options, as well as a lack of cultural humility among clinicians(16,17). This creates an escalating cycle of need when help-seeking is met with dismissal, blame, or failures to understand contextual realities, which erodes trust and has ripple effects for future help-seeking for the individual and their networks(18).

Where policy fails to address these inequities or ensure effective implementation strategies translate equity-oriented frameworks into practice(19,20) impacted communities are driven to seek and create supports that understand and align with their needs. That is, community-led services may not only offer located expertise or potentially ‘safer’ spaces for marginalised communities but also respond to gaps in the public system, providing vital preventative and crisis-responsive supports. Given the history of institutionalisation in mental health systems, deeply intertwined with ableism, racism and classism in England, keeping concerns hidden, *within* trusted networks and out of state systems may for some feel essential to protection and maintaining agency.

### The conditions for community care

However, communities are also enabled – and constrained – in providing care; what services are able to offer is shaped by access to resources, perceptions of valid expertise, and political climate. Many community-led services depend on grants or charitable donations, producing funding instability that impedes staff retention, training, and investment, restricts long-term planning and risks the availability, continuity and quality of provision – all while intensifying the practical and emotional demands on providers (21,22). Choice, changes in or even loss of provision can have detrimental impacts, disrupting continuity in care and compromising service engagement(23). Funding allocations are often shaped by the same forces that govern public services. Increasingly polarised attitudes toward diversity, equity and inclusion initiatives alongside a fiscal context of austerity, that has fallen heaviest on marginalised communities (24), leave services supporting these communities particularly vulnerable while navigating increasing demand. Inequities also arise at the point of obtaining funding, where the expertise of bidding and reporting - distinct from community or care expertise – is rewarded, advantaging well-resourced organisations. As such, understanding the community care landscape requires attending to not just what exists but the conditions under which these services operate.

### What is known about community services in England?

Previous mappings of mental health support for children and young people in England have established that provision is diverse, with just under half of services provided by the third sector (23). Targeted ‘LGBTQ+’ and ‘ethnic minority’ supports for young people were provided predominantly by voluntary/community organisations, rather than the NHS and concentrated in urban and more ethnically diverse areas, indicating an implicit reliance on the sector to provide specialised mental health provision for minoritised young people (25,26). While these mappings provide valuable national-level insights into service variation, by primarily collecting data from formal bodies (NHS services, Integrated Care Boards, Local Authorities), with supplementary internet searches, or use of select online service directories (23,25) they offer limited insight into community-based supports outside of formalised or commissioned services. These online databases, still evolving, capture only a selection of community-based services, and lack information on specific equity-oriented frameworks. Without explicitly examining equity-oriented approaches, questions remain around the scope, inclusivity and accessibility of these services. Additionally, broad-scale mappings cannot capture the nuances of localised community care landscapes.

### Rationale & aims

Guided by the principles of community-based participatory action research (27), we aimed to address these knowledge gaps through an equity-oriented mapping of place-based community services in London and Greater Manchester supporting children, young people and family mental health and wellbeing. Though communities transcend localities, given that delivery mode shapes access, provision and operational realities as well as how inclusion, exclusion and safety are experienced, we focus on services facilitating physical coming together. Recognising that in the socio-political-historical contexts of systemic oppression and exclusion, different groups need different care to achieve good mental health or to attend to their wellbeing in ways that make sense for them, we sought to understand the specificity of equity-oriented practices. By focusing on non-commissioned services and integrating expertise of community stakeholders, we explored the contextual realities of community care outside of the state system, the needs these services are attending to, as well as remaining gaps and ongoing barriers to provision.

## Methods

### Conceptual Framework

This approach was guided by the principles of Participatory Action Research(27), which emphasises meaningful partnership between researchers and people with lived experience of or those most affected by a research topic. Crucially, attending to and confronting power dynamics with the goal of redistributing power equally across all voices(28). Our lens is rooted in Black feminist scholarship, particularly intersectionality (29), recognising the impacts of complex, interacting oppressions, emphasizing the necessity of systemic transformation. We align with the tenets of transformative justice, using knowledge to drive systemic change, amplifying community efforts, imagining alternative realities and proactively working towards them (30). We are intentional about centring wellbeing and care, integrating trauma-informed (31) and cultural safety (32) principles in an effort to minimise harm, foster reflection, and redistribute power. Our working together and with stakeholders has also been informed by Indigenous approaches to experience and knowledge sharing, emphasising respect, reverence, responsibility and reciprocity (33).

### Co-production Approach

This project was co-produced with three young people with lived experience (co-researchers) and community stakeholders (policy advisors, mental health professionals, special educational needs and disability co-ordinator, community providers, youth workers and parent/carers) who were recruited through community-based organisations, community networks, word-of-mouth and direct outreach as part of a larger project examining intersectional discrimination in pathways to mental health care for children and young people in England.

Team discussions guided conceptualisation, question prioritisation, and protocol development. Co-researchers engaged in collective decision-making throughout each stage, each opting-in to a lead role for different aspects aligned with their interests and goals. One contributed to protocol materials and survey development, leading copy editing and creative tasks as well as training in coding and analyses. Two trained in data collection, extraction and verification; one led searches of resource and signposting lists, another led website verification and geographic mapping. A stakeholder with expertise in youth work and mental health research also trained in and completed data collection. All co-researchers and stakeholders contributed to the analyses, interpretation, write-up and recommendation development.

Participation began with co-developing our ways of working and investing in relationship-building. This foundation established open communication, attunement and flexibility to needs, enabling co-creation and allowing for continuous growth. Co-researchers share reflections on co-producing this analysis to provide insight into our ways of working and the meaning of embedding lived expertise across the research cycle. Reflections are presented anonymously, in accordance with our working together practices which centre trauma-informed and cultural safety principles, ensuring recognition as co-researchers and co-authors without expectation of disclosure.

Our approach to participation felt intentional, starting with treating all expertise as equal, and shared decision-making as a core value, shaped the ways we worked together. Working on the service mapping felt meaningful, not just because of the tasks we completed, the progress we made or even the findings, but because of how we worked together. Pulling together different experiences, creating opportunities for brainstorming and sharing ideas, and taking the time to reflect on what was working or not working, enabled us to succeed, build confidence and become closer as a team. During the data collection, it was interesting to see the number of potentially ‘eligible’ services available, and to be a part of compiling them - even the ones that did not meet criteria for our analyses. It felt like we were working on something in service of communities, not just research.

At the same time, working on this project was time-consuming. Because the data collection and verification processes were repetitive, at times, it was hard to maintain motivation. The sheer number of lists and services also meant during the process we lost sight of the scope of the work and needed to actively come back to the goals of the project. Accessibility issues and unclear websites made it more difficult to make decisions and feel confident in them, slowing the process further. At times, it felt draining reviewing multiple services as this highlighted the extent of unmet need. Working in the background, on ‘back end’ tasks, meant it was easy to lose sight of the impact of standalone tasks completed in shorter time frames even if they were significant contributions. However, having a supportive team, cheering each other on and being open to problem-solving together meant that we were always able to find ways forward.

Our commitment to flexibility and choice was crucial. Throughout the project there were times when we chose to opt-in or opt-out of leading or contributing to different components. As a team we are all very understanding and know that sometimes people have different energy levels. When scheduling, this meant understanding that availability and capacity fluctuate, making sure everyone knows it’s okay to take a break or press pause on the work if they need to, and finding ways to catch-up or contribute at a pace that suits. In those moments, instead of saying that we’ll all push through, people with more energy and headspace took on more work than those who had low energy. This was particularly important as we recognised the different responsibilities and demands that everyone was juggling.

*Ultimately, reflecting on our work, seeing the tangible results of the project was rewarding, and the opportunity to apply our expertise in a meaningful way was fulfilling*.

### Mapping Methodology

#### Mapping Settings

Service mapping was conducted across London and Greater Manchester. London is the biggest city and considered the most diverse region in England, with an estimated 63.2% of residents from racially and ethnically minoritised backgrounds. In 2021, 22.7% of London’s population was aged under 18(34). Greater Manchester is the second most populous urban region, with 28.7% of Greater Manchester’s residents from racially and ethnically minoritised backgrounds, with a higher proportion among children and young people (34.0%) (35). Manchester City Local Authority is considered the most diverse district in the north of England, with 51.3% ethnically minoritised residents(36).

Although both regions are ethnically diverse, with growing awareness of the need for cultural sensitivity and equity-oriented approaches, the demographic composition and policy contexts differ. Greater Manchester has implemented the THRIVE framework (37), including Thrive Hubs as single-entry points to access advice, signposting and service navigation. Both cities have also experienced instances of collective trauma impacting young people and families in recent years, for example, the Manchester Arena bombing (2017) and Grenfell Tower fire (2017), heightening awareness of trauma and mental health needs in affected communities (38,39). This context made these settings particularly relevant for examining equity-oriented community-based provision.

#### Eligibility criteria

We defined our target service criteria as (1) community-based (2) not Government, National Health Service or Local Authority provided or commissioned (3) providers of mental health or wellbeing support (4) for children aged 0-18 years or parents, carers, families, kinship in relation to those under 18 years in their care (5) currently operating with options available free of charge. We purposefully took a broad approach to ‘support’, defining this as services that self-identified as supporting mental health, emotional development, behavioural regulation or promoting emotional wellbeing ([Supplementary Table 1 Keywords](https://liveuclac.sharepoint.com/:w:/r/sites/PeerResearchTeam/Shared%20Documents/Service%20Mapping/Project%20Manuscript,%20Report%20%26%20Presentations/Manuscript/Community%20Service%20Mapping%20Paper/Supplementary_Files.docx?d=wd2467c975eeb4272bf9086ebec5a749e&csf=1&web=1&e=rX3bJG)).

Services were excluded if they were: only delivered online, through schools, or in hospitals; government provided or commissioned/contracted; referred only to ‘health’ without reference to wellbeing, emotional or mental health-related keywords; not in relation to 0-18 year olds; only available fee-for-service.

#### Sources & Search Strategy

Our initial protocol built on Price and colleagues (40)’s seven steps to mapping health service provision but adapted for community services, to overcome additional barriers to survey-based data collection. Pilot scoping (October/November 2023) informed refinement of our scoping strategy and eligibility criteria.

*Survey.* In November 2023, we launched a brief survey to gather information from support-seekers, providers and other stakeholders. This was distributed to key informants and organisations, targeted outreach, as well as through national and community-based organisations’ networks (including promotion by the Association for Child and Adolescent Mental Health, the Children and Young People’s Mental Health Coalition newsletter, and a co-written commentary on the Race & Health website). Despite these dissemination efforts, survey recruitment proved challenging and yielded limited responses, leading us to concentrate our efforts on the systematic search approach.

*Searches.* Our search strategy employed both systematic and snowball sampling approaches to capture services that may not be listed in formal directories. We began with searches of established signposting platforms, resource lists and databases (including, Hub of Hope, Bayo, Best for You, Joy, Youth Work One). We adopted a snowball approach, systematically following all links to additional services and signposting resources until no new links or potentially eligible services were identified. Additionally, we systematically searched Local Authority ‘Local Offer’ websites and NHS resource lists for integrated care board areas.

Data collection continued November 2023 through July 2024, followed by an update and verification period (January-March 2025). The mapping period closed on 31 March 2025.

#### Screening & Verification

Services deemed ‘potentially eligible’ were collated and cross-checked for eligibility against our eligibility criteria. All entries completed by team members were subsequently cross-checked by project lead to ensure consistency. Regular team meetings enabled collective decision-making regarding borderline cases, co-working sessions and check-ins supported ongoing alignment in interpretation of eligibility criteria. Where eligibility could not be determined due to lack of publicly available information online, invalid or inactive website, they were checked a minimum of three times over a 1-month period before being categorised as unverifiable.

Where eligibility depended on funding status, we obtained financial histories from the [Charity Commission for England](https://register-of-charities.charitycommission.gov.uk/en/)(41)using the most recent annual return available and publicly available grants data from [360GrantNav](https://grantnav.threesixtygiving.org/)(42); services reported to be in contract with or commissioned by government were excluded. Services funded by government grants, but not contracts, were deemed to be sufficiently independent governance. For Community Interest Companies not registered with the Charity Commission, we requested funding information directly from services. Where services did not respond to verification requests, eligibility determination was based on publicly available information only.

We then extracted publicly available information of ‘eligible’ services and contacted them by email seeking to confirm the accuracy of the publicly available details we had collected. Additionally, we invited services to share additional information and insights, via email, phone-call or consultation to discuss further.

We present a snapshot of services as verified between January-March 2025. Services that were operating when initially identified but closed during the mapping period as well as those that entered into a contract with government were excluded during final verification.

#### Analyses

We extracted and synthesised data on service characteristics including: geographic location, target age groups, specialisation, referral/access requirements, types of provision, and equity-oriented components including: (i) cultural safety, anti-racist or culturally specific approaches, (ii) trauma-informed or trauma-sensitive frameworks, (iii) language options or translation, (iv) immigration-related support or accessible regardless of immigration status, (v) explicitly intersectional orientation, (vi) LGBTQAI+ inclusive or specific, (vii) faith-based or specific, (viii) gender-specific, (ix) special educational needs and disability inclusive (Supplementary Table 2).

We completed a narrative synthesis of themes and contextualisation of findings. During the screening and verification processes, we adopted a “support seeker lens”, considering what information would be available and comprehensible to a young person or family seeking support and noting observations about accessibility of information, clarity of provision, and gaps or inconsistencies in what was communicated. In conversation with stakeholders and integrating insights shared by service providers during the verification process through email correspondence (16), brief chats (2) and in-depth consultations (3), this interpretive analysis examined patterns with respect to our research questions, considering the meaning and implications of findings for accessibility, equity, and the broader landscape of community care.

## Results

### Sources

The iterative process involved 118 sources, including existing databases, resource lists and service navigation signposting, local authority ‘Local Offer’ websites, community organisation partnership listings, targeted internet searches of boroughs, survey responses and word-of-mouth recommendations from team members and stakeholders. A complete list of sources is available in Supplementary Table 3.

The scoping process yielded 1110 ‘on first glace’ potentially eligible services after duplicates were removed (Figure 1; Supplementary Table 4). After cross-checking against our eligibility criteria, we identified 297 community-based services advertising active support for children, young people and family mental health or wellbeing not reported as commissioned by or in contract with government according to publicly available records for the financial year 2024. 51 services were deemed unverifiable due to insufficient publicly available information i.e. no or inactive website. Reasons for ineligibility included lacking explicit advertisement of active mental health or wellbeing-related support for children, young people, parents or families, being government or NHS commissioned/contracted, being private fee-for service only with no free/subsidised offerings, not being community-based (e.g. only available online or through schools), out-of-area or resources and signposting only. Just under half (48.9%) of excluded services were found to be contracted, commissioned or delivered by government agencies.

**Figure 1.**
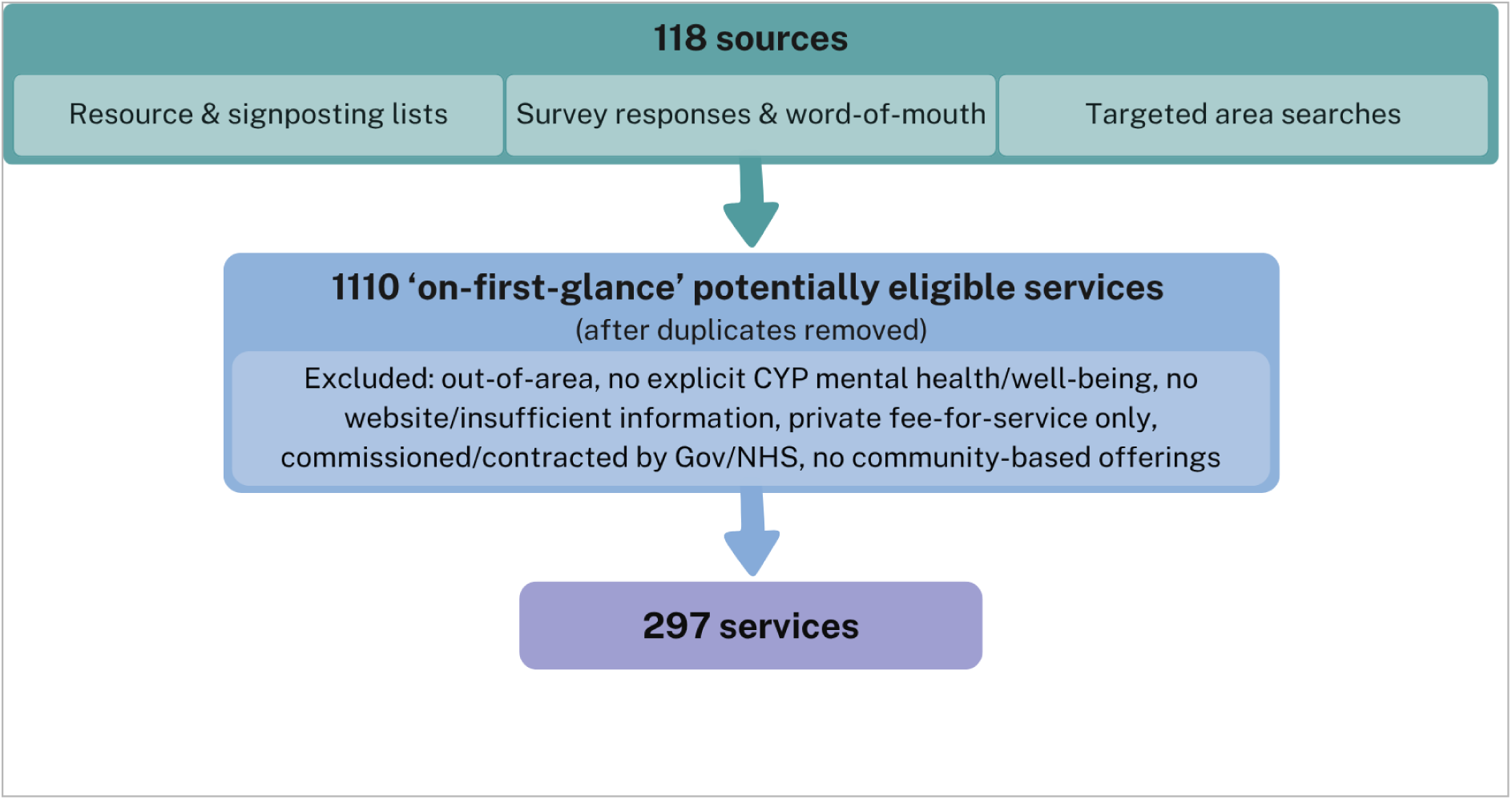
Scoping Services Flowchart.

### Geographic Mapping

Of the 297 services, 261 were based across London and 41 were across Greater Manchester, with 5 listed in both (Supplementary Table 5 & Supplementary Table 6). We report overarching trends across all included services, specifying where trends differ substantially across locations.

Broadly, boroughs with higher ethnic diversity and deprivation had higher numbers of community-based services (e.g. Lambeth, Southwark, Hackney, Tower Hamlets, Newham, Camden, Brent), however, this was not a consistent pattern (e.g. Waltham Forest). Lambeth stood out with the highest number of services (31), a borough with a long history of well-documented structural inequities and strong traditions of Black-led community organising.

### Scope & Access

Services appeared primarily targeted toward older children and young people, with over half of services advertising support for those aged 11 years and older, least likely to provide early intervention support for children under 5 years, with 24% of services not specifying an age range (Figure 2a).

**Figure 2.**
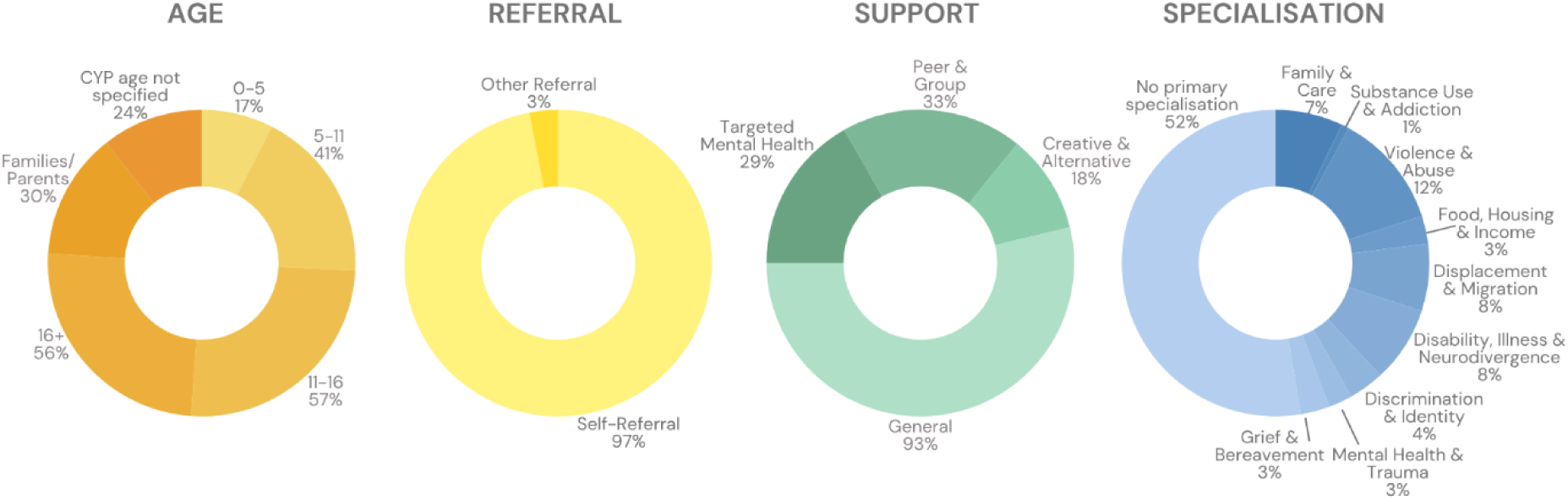
Proportion of services (a) providing services for different age ranges* (b) referral requirements (c) offering different types of support* (d) identifying with a primary specialisation. *Note: Many services provided services across age groups and offered multiple types of support adding up to >100%.

Nearly all services (97%) accepted self-referrals, either from parents, caregivers, family members, or - if over 16 years - directly from the individual seeking support (Figure 2b). Three percent of services required a referral through another organisation.

Most services (93%) provided general non-specialised wellbeing support, including mentoring, youth work, sport, psychoeducation and practical support (Figure 2c). One-third provided targeted peer or group support (33%) with slightly less offering specified targeted mental health support (29%), such as counselling, talking therapy or a therapeutic program. Fewer (18%) advertised creative and alternative supports, such as art, music, drama, animal or holistic therapies.

Approximately half of services (52%) did not specify a primary area of specialisation (Figure 2d). Among those that did, violence and abuse support (12%) was the most frequently offered service, covering both community-based and interpersonal violence. Support specialising in substance use or addiction was relatively absent (1%).

### Equity-Oriented Provision

Overall, 84% of services advertised group-specific or equity-oriented provision (London 85%, Manchester 73%) (Table 1). Practices related to cultural safety, anti-racism, or providing culture, race, or ethnicity specific services were most frequent (38%), followed by ‘accessible’ services or inclusive of individuals with special educational needs and disabilities (37%). While 29% of Manchester services advertised supports for specific gender identities, this was 20% in London. In contrast, 13% of services in London identified as faith-based or faith-specific, this was 5% in Manchester. Just 11% of services overall (17% Manchester, 11% London) explicitly adopted an intersectional lens or targeted multiply minoritised individuals.

**Table 1.** Equity-oriented provision.

|  | Cultural safety or specific | Trauma-informed | Language | Migration status | Intersectional | LGBTQAI+ | Faith | Gender | Disability or accessible | Any equity-oriented provision |
| --- | --- | --- | --- | --- | --- | --- | --- | --- | --- | --- |
| **Manchester** | 34.1 | 12.2 | 26.8 | 29.3 | 17.1 | 14.6 | 4.9 | 29.3 | 36.6 | 73.2 |
| **London** | 38.3 | 21.8 | 21.8 | 29.9 | 10.7 | 16.9 | 13.4 | 20.3 | 37.2 | 84.7 |
| **Overall** | 38.0 | 20.9 | 22.6 | 30.0 | 11.1 | 16.2 | 12.5 | 21.5 | 36.7 | 83.5 |

Of services that advertised any equity-oriented provision, 34% advertised just one, 21% advertised two, while 45% advertised three or more (e.g. cultural safety, trauma-informed and immigration status). Services advertising multiple equity-oriented approaches clustered most frequently at intersections of cultural safety or specific care, migration status, language support and trauma-informed practice. Less frequent intersections were found between faith-sensitive, LGBTQAI+ and gender specific supports (Figure 3).

**Figure 3.**
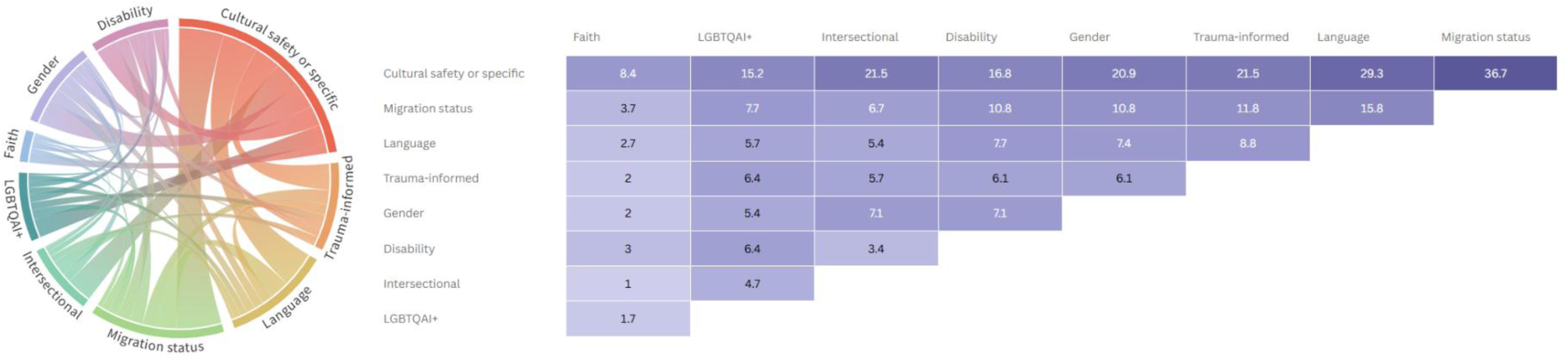
Intersections of equity-oriented provision. (left) Thickness of threads between practices representing the proportion of services who aligned with both practices. (right) Cells represent the proportion of services who aligned with both practices.

Among services with an identified specialisation, those focusing on violence and abuse, displacement and migration, as well as discrimination and identity-specific care were most likely to advertise equity-oriented provisions (Table 2). Notable gaps were identified, particularly for services specialising in supporting children and young people with disabilities, illness or neurodivergence, while providing accessible or disability inclusive support, there was minimal integration of other equity dimensions. Faith-based or sensitive support was consistently low across all specialisations and explicitly intersectional approaches were rare beyond services specialising in violence and abuse and discrimination.

**Table 2.**
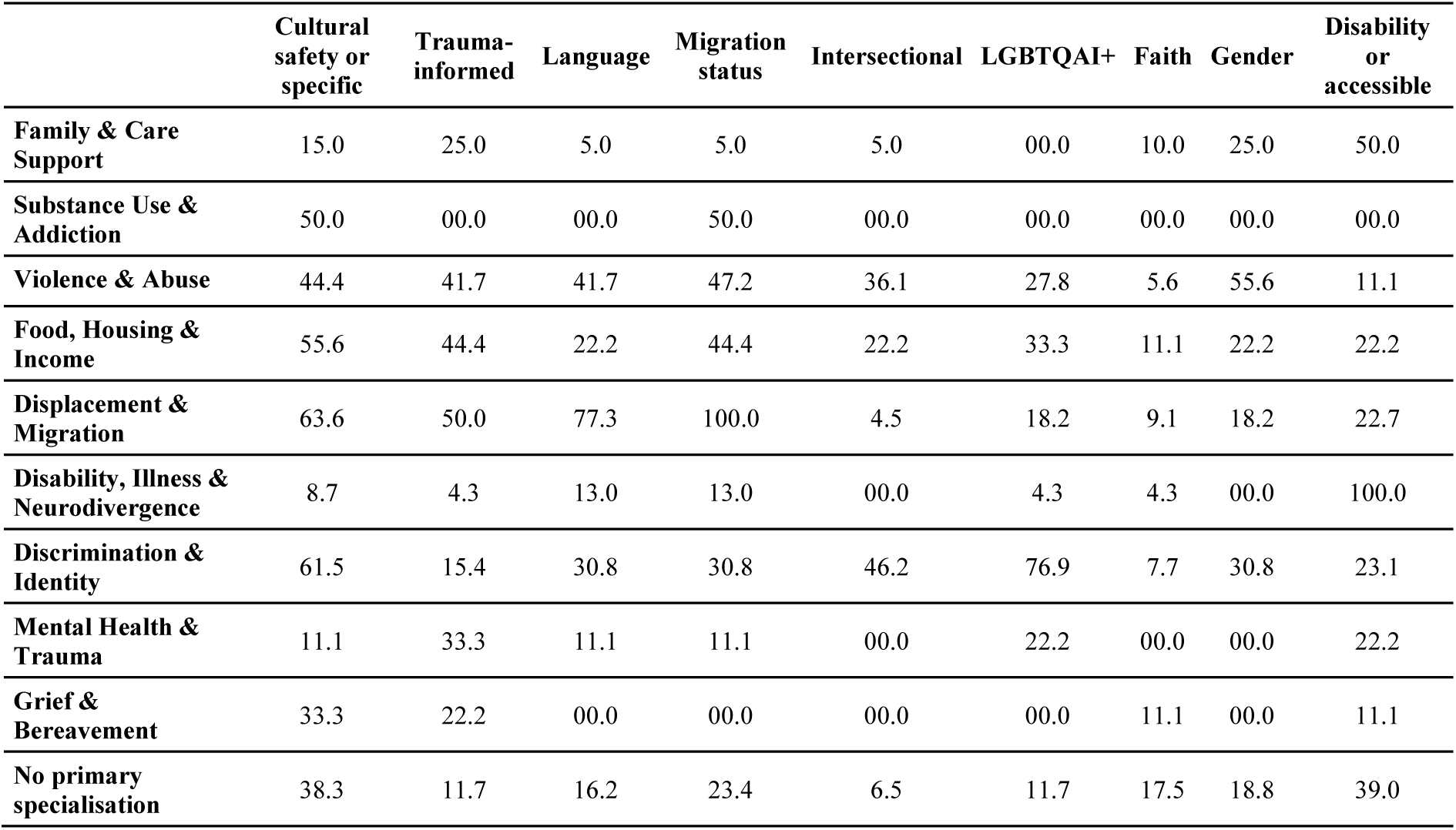
Equity-oriented provision by Primary Specialisations.

|  | Cultural safety or specific | Trauma-informed | Language | Migration status | Intersectional | LGBTQAI+ | Faith | Gender | Disability or accessible |
| --- | --- | --- | --- | --- | --- | --- | --- | --- | --- |
| **Family & Care Support** | 15.0 | 25.0 | 5.0 | 5.0 | 5.0 | 00.0 | 10.0 | 25.0 | 50.0 |
| **Substance Use & Addiction** | 50.0 | 00.0 | 00.0 | 50.0 | 00.0 | 00.0 | 00.0 | 00.0 | 00.0 |
| **Violence & Abuse** | 44.4 | 41.7 | 41.7 | 47.2 | 36.1 | 27.8 | 5.6 | 55.6 | 11.1 |
| **Food, Housing & Income** | 55.6 | 44.4 | 22.2 | 44.4 | 22.2 | 33.3 | 11.1 | 22.2 | 22.2 |
| **Displacement & Migration** | 63.6 | 50.0 | 77.3 | 100.0 | 4.5 | 18.2 | 9.1 | 18.2 | 22.7 |
| **Disability, Illness & Neurodivergence** | 8.7 | 4.3 | 13.0 | 13.0 | 00.0 | 4.3 | 4.3 | 00.0 | 100.0 |
| **Discrimination & Identity** | 61.5 | 15.4 | 30.8 | 30.8 | 46.2 | 76.9 | 7.7 | 30.8 | 23.1 |
| **Mental Health & Trauma** | 11.1 | 33.3 | 11.1 | 11.1 | 00.0 | 22.2 | 00.0 | 00.0 | 22.2 |
| **Grief & Bereavement** | 33.3 | 22.2 | 00.0 | 00.0 | 00.0 | 00.0 | 11.1 | 00.0 | 11.1 |
| **No primary specialisation** | 38.3 | 11.7 | 16.2 | 23.4 | 6.5 | 11.7 | 17.5 | 18.8 | 39.0 |

### The Contextual Realities of Community Care

Through our collaborative analyses we identified a series of trade-offs and tensions faced by community service providers, drawing on observations from our scoping process alongside insights shared by stakeholders and services.

#### Responsive but under-resourced

The extent of public sector reliance on third sector provision was evident with half (48.9%) of potentially eligible services commissioned/contracted while many included services relied on public sector grant funding. Given the majority aligned with at least one equity-oriented practice and at times born organically in response to community events or trauma (e.g. Kids on the Green emerging in response to the Grenfell Tragedy), community-based services appear highly attuned to the needs and lived realities of the populations they serve, continuously evolving alongside and in response to changes in community needs. Often offering multiple types of supports, they accommodate different needs at the same time and allow for individual needs or preferences to change while maintaining a sense of stability and continuity in support. However, services founded in response to crisis prioritise immediate delivery over strategic business planning, or ensuring practices are evidence based and sustainable long-term. That is, responding to immediate need sits in tension with the time and resources required to ensure all providers are trained in specific practices.

#### Provision versus promotion

While there was considerable overlap across resource lists and signposting services, with each additional list we continued to unearth new ‘potentially eligible’ services. Additionally, the scoping process indicated wide variation in the availability of services as well as service navigation tools; While some local areas had comprehensive resource lists and a greater range of ‘potentially eligible’ services even with borough-specific searches finding publicly available information on community-based services in certain locations was challenging. Service deserts do not inherently indicate more or less need, but may reflect variation in priorities and funding models across local areas.

Once identified, service websites were often inaccessible or outdated, difficult to navigate, or lacked basic information, making it challenging to understand the scope and types of support offered. Information on eligibility and suitability criteria, therapeutic approaches or training of providers was largely absent. Although specialised services often identified their target population, very few services explicitly communicated their limitations – that is, who they were not resourced to support or what would be out of their scope of practice due to training or ethical considerations. Hesitancies to disclose limitations related to confusion around Equality Act adherence, and fears of misinterpretation, reputational risk or that disclosed limitations could be weaponised against them.

#### Strategic (in)visibility

Despite the abundance of, albeit fragmented, resource and signposting tools, it is likely many services remained effectively ‘hidden’ - reflecting practical constraints as well as intentional choice. Prioritisation of provision over marketing, limited resources and insufficient capacity to develop and maintain a website and stop-start provision due to short-term funding cycles all impede promotion. Others may actively choose to stay hyperlocal, in local area or identity community, based on word-of-mouth or limit publicly available information as part of safeguarding procedures protecting support seekers. Additionally, informal, ad-hoc support through networks, faith-based organisations or places of worship is often not framed in mental health or wellbeing terms, operating instead through unwritten understandings between community members rather than publicly advertised provision. Stakeholders noted even within networks there may be strategically invisible community networks, selective for safety or fear of scarcity in the supports they have come to rely on.

Similarly, through the verification process, a gap emerged between what equity-oriented practices services publicly advertised and approaches they were trained in or identified with through private channels of communication. What services made publicly visible appeared influenced by overlapping considerations including, community perceptions, fears that by advertising some inclusive practices this might inadvertently exclude or unintentionally discourage others, as well as updating publicly available information falling to the bottom of the to-do list. For some, selective communication was viewed as a ‘protective measure’ for both services and those seeking support to manage expectations and avoid misinterpretation. Services expressed concerns around advertising inclusive practices that they may not have capacity to sustain if faced with funding cuts, high demand, staff leave or absences. Services also noted website content functions more as a portfolio for funders than a public outreach tool.

#### Intention, expectation or box-ticking?

At times there appeared to be a disconnect between identified needs and approach, for example, services providing support for “trauma”, without using trauma-informed, sensitive, or responsive approaches. Many services mentioned having predominantly racially or ethnically minoritised attendees, often using the reductive term “BAME”, yet did not advertise practices grounded in cultural safety, anti-racism, or anti-oppressive frameworks. Similarly, some services advertised or during verification reiterated a stance that they “do not discriminate” though did not identify with anti-discrimination frameworks or practices. Services also referred to “SEND” without defining who this includes, or what needs they can meet, recognising the wide spectrum of presentations, multiple co-occurring needs and environmental or sensory accessibility considerations that fall under the umbrella term. For some, naming the needs or demographics of their communities may reflect good intention but without sufficient training, resourcing, or capacity to implement required practices – or without the knowledge or shared language to reflect their work embodies these practices. For others, these gaps may reflect unintentional oversight, genuine confusion about what equitable practice requires, or overly cautious interpretations of the Equality Act. For others still, the use of keywords represents a tool to access funding, driven by funder expectations and priorities, to appear relevant and maximise chances of obtaining funding.

#### Structural realities shaping funding precarity

During the mapping period, we observed services closing due to funding cuts and services both entering and exiting government contracts. Short funding cycles were characterised by insufficient time to demonstrate impact, plan strategically, embed consistent models of support, recruit, retain or build expertise. Once-off or short-term projects were critiqued as failing to account for the realities of service delivery - for slow starts, learning curves, time required to hire, train, advertise and build trust, demonstrate meaningful impact - or effective supports for multiple, co-occurring or long-standing needs. Additionally, expectations of immediacy and impact measures focused on crude access numbers, rather than the experiences of those accessing or the gap being filled, risk attendance inflation and cycles of funding ineffective but seemingly ‘well-attended’ services.

‘Calls for bids’ were described as influenced by individual commissioners or funder priorities, often reactive to incidents, public reports or political agenda rather than developed in consultation with communities or providers. Services noted that in the absence of avenues to fund ongoing prevention or early intervention projects, they feel forced to ‘fit in’ to reactive funding calls. Anxieties were amplified by concerns around political determination of funding allocation - government priorities, political climate and shifting public attitudes toward equity, diversity, and inclusion - leaving services supporting minoritised and marginalised communities particularly vulnerable, especially based on racial-ethnic group, nationality, migration status, religion, gender identity, or sexual orientation.

#### Unaffordable choices

Trade-offs emerged between commissioning contracts and grants. While grants offer greater flexibility and less extensive reporting requirements, allowing services to provide rapid response and pivot based on community need or feedback, contracts offer greater financial security if renewed or extended and establish clear remits or may facilitate relationships with statutory bodies. At the same time, contract parameters may restrict flexibility to adapt to emerging community needs and impose data collection and sharing requirements, without guidance or support, that demand time, capacity and specialist skillsets services may lack. However, access to funding was often shaped by necessity rather than choice, reflecting inequities in funding processes amid high demand in a context of constrained public funding and fractured political climate.

Obtaining funding also requires specific skillsets - grant development, knowledge of ‘key words’ and report writing expertise – distinct from expertise required for direct provision, or even operational management. Grassroots organisations are forced to compete with well-established organisations that have employees or teams dedicated to grant-writing, longstanding relationships and prior government contracts, who dominate funding allocations, exemplified in the mapping process with many seemingly smaller services provided by large organisations with established contracts (e.g. CGL, Family Action, Catch22). Competition and time-pressures create risk as services may overstate previous experiences or over-promise what they can deliver within unrealistic timeframes – driven by good intentions or survival instincts.

#### Defining independence

Given that funding is constantly in flux, so too are funder requirements and data-sharing agreements. This complicates perceptions of independence which may be understood differently by service providers and community members. We found a noticeable lack of clarity around funding sources and the role of public authorities in service provision, in some instances, services advertised themselves as “independent” while contractual ties suggested a more bound relationship. For services, intermittent or partial commissioning may not feel like a bound relationship, offering no more stability or security than a once-off grant. However, public perceptions of ‘independence’ may be grounded in the assumption of no government involvement, revealing how differently ‘independence’ is understood by services and by the communities they serve.

## Discussion

### Summary

We identified 297 community-based services across London and Greater Manchester supporting the mental health and emotional wellbeing of children, young people and families, the majority of which aligned with at least one equity-oriented practice (84%). Almost all (93%) services provided general wellbeing support either solely or alongside targeted or alternative supports, predominantly serving older children and young people. While most services were open access, a handful of services (3%) required referral through other statutory organisations, meaning families needed to be in contact with and to have disclosed need to government-provided services. Just under half identified a primary specialisation, broadly, these specialised services had higher equity-oriented provision, often matching practices with populations served (e.g. trauma-informed violence/abuse support or displacement/migration support offering multiple languages). Services supporting those who have experienced violence and abuse, as well as those navigating displacement and migration, most explicitly aligned with equity-oriented provision, including intersectional supports. While most services did not explicitly advertise attending to multiple, intersecting forms of discrimination, a small minority did.

Our collaborative analysis indicated a responsiveness and need-driven evolution of services – filling critical gaps in statutory services – although their agility and prioritisation of immediate need brings challenges for sustainability, capacity-building and adoption of evidence-based practices. The scoping and verification processes highlighted the fragmented and challenging nature of service navigation alongside conflicting priorities and cautious information sharing which act as barriers to accessing the right support even if available. Broad umbrella terms (e.g. BAME, SEND) may intend inclusivity but risk homogenising diverse experiences and appearing tokenistic, while assertions that services accept difference or uphold Equality Act duties (43) imply the same colour-evasive approach statutory services are critiqued for. Throughout, the pervasive implications of funding precarity were evident alongside the political determination of and inequities obtaining funding, and implications for perceptions of government involvement and data sharing requirements. These structural realities shape what services are deemed fundable, and in turn, what support is provided, to whom, or whether services can afford to offer anything meaningful at all.

### A (precarious) lifeline

The reliance on community-based services to fill gaps in statutory provision was evident throughout our analysis. Community services may be uniquely positioned to respond to community needs and offer aspects of safety through cultural grounding, shared understanding of experience, and relational trust that public systems have failed to adequately invest in. However, they face similar challenges to statutory services related to scope of practice (the safe boundaries based on education, competence and professional regulation), and potential contradictions between well-intentioned but harmful practices - yet in the absence of stable resourcing and organisational safeguards. Additionally, constraints impede access to training or learning opportunities to address knowledge gaps, particularly if services are faced with a trade-off in time and resource between provision and training. Ultimately, the constant funding precarity creates risk and instability for services and those they serve. This is not just directly through the loss of valuable support (44) but also unintentional indirect risks as services may find themselves operating without adequate training, resources or infrastructure, struggling to maintain continuity in care, cutting corners or providing potentially ineffective practices.

### Enabling informed choice

Choice is a central component of trauma-informed systems(31), however, our findings indicate a number of conflicting tensions around transparency alongside previously documented access barriers(23). Fragmented signposting and lack of publicly available information, alongside knowledge gaps between the public system and specialised community services(25) create uneven availability and awareness. Services face legitimate concerns about misunderstanding, miscommunication or over-promising, however, insufficient information sharing risks undermining access to appropriate care through unsuitable referrals, false-starts, ineffective mismatches and unrealistic expectations of the support available. In turn, those with greater system literacy, resources or in specific areas, who have the time and knowledge to locate or contact services, as well as the skills and knowledge to ask the ‘right’ questions, have greater access, risking widening inequalities. Similar to the criticisms of CAMHS, hidden eligibility criteria and case-by-case decision-making, leaves room for discrimination or differential treatment, while transparent communication informs support seekers prior to referral, reduces administrative burden related to unsuitable inquiries and reduces potential deterrence that comes with rejection. This does not mean overwhelming audiences with information but ensuring sufficient information is available to make an informed decision about whether or not the service is available to them (eligibility), what the service can and cannot provide (suitability), and the values, ethos or cultural orientation (framing) allowing families to ascertain if it is a good match for their needs. Open communication not only enables informed decision-making(45) and builds trust through candour about what a service can and cannot do(46), but also makes visible those children and young people falling through gaps or being bounced between services.

### Who is still being missed

While community-based services are filling crucial gaps in the public system, there remain gaps in provision, meaning some communities are neglected or faced with trade-offs in their pursuit of care. The wide variation across boroughs – in services and navigation tools – aligns with well-documented geographic disparities meaning families living in service deserts face additional barriers(25). As mapped previously services more frequently supported secondary school age children(23,25), corroborating the need for more emotional wellbeing prevention and early intervention supports for young children and parents in addition to early learning investments(47), particularly for families more at risk of experiencing birth-related trauma due to health system failures(48). Given the higher proportion of services supporting those who have experienced violence and abuse, and the government’s mission to halve violence against women and girls(49), there is a clear need for preventative, early intervention, transformative justice approaches to interrupt cycles of trauma, prevent future violence and reduce high need for later crisis response.

Crucially, services explicitly meeting the needs of multiply minoritised communities remain limited. This risks families feeling forced to choose between aspects of their identity or needs or that they have no choices at all(44). For example, given services with specialisms in disability, illness or neurodivergence were less likely to advertise equity-oriented provisions, a racially minoritised family seeking wellbeing support for their child with a disability is faced with the reality the service might be resourced to support the disability but may not be suitable to understand the cultural context or may be culturally unsafe. At the same time, culturally specific services may understand the socio-cultural contexts of communities and seek to respond but be ill-equipped to appropriately support a child with multiple co-occurring needs. This highlights a need for cross-sector knowledge and skill-sharing, to enable services to attend to the needs of multiply minoritised families through accessing peer supervision and clinical consultation and facilitate a bridge to specialised supports if needed.

### The need to bridge the gap

While the Patient and Carer Race Equality Framework (PCREF) for NHS mental health trusts became mandatory from March 2025(50), implementation remains in early stages with scarce reporting in CAMHS. Instead, the attitudinal stance ‘everyone is treated equally’ – prevalent in mainstream service provision – implies a ‘colour-evasive’ approach(51), which risks reproducing power dynamics and perpetuating discrimination. Similarly, there is limited guidance on trauma-informed approaches in policies related to children’s needs (19) and no national strategy or funding commitment (52) despite endorsement in adult mental health policy. This suggests that the organisational change required for equity-oriented statutory services remains unrealised – though expertise community providers hold.

Statutory services are often positioned as the ‘expert’ or default ‘safe’ option – however, this risks elevating clinical or government expertise above that of communities and fails to recognise these services may not hold relevant expertise or feel safe for everyone. The reputation that accompanies government-commissioned services may for some instil a sense of trust but for others, hesitancy or anticipation of harm. However, limited accountability mechanisms in community services also raise safeguarding concerns. The same invisibility that offers protection for communities accessing hidden supports can also place them beyond safeguarding oversight or mechanisms to ensure safe practices.

In line with the national youth strategy (53), shifting towards greater collaboration between statutory and community services has the potential to support expertise sharing and capacity building between the sectors - both for community-based services and for statutory service providers. Recognising these services serve different needs, hold different areas of expertise, offer different specialisms, there is an opportunity to fill gaps and work together in complementary ways with the potential to act as a ‘bridge’ meeting communities where they are while also supporting access to statutory services when needed and when families are ready.

For statutory services, given understanding community needs and contextual realities as well as addressing implicit bias and discrimination is an ongoing commitment(54,55), investing in longer-term community consultations, rather than once-off trainings, embeds tangible – and reciprocal – engagement that facilitates (cross-system and therapeutic) relationships, trust-building and co-ordinated supports. For community services, with constrained resources, partnership working with CAMHS, consulting on children of concern and co-working with children and families, has the potential to build capacity, facilitate more appropriate referrals and supports, and embed an additional safety net, particularly around safeguarding, evidence-based practice and risk management. This is not to be mistaken for ‘policing’ services, imposing judgement of or directives on how to care but rather to establish safe boundaries around different areas of expertise and ensure a wider eco-system of care. Alongside greater transparency around working together practices, government or statutory service involvement, these practices have the potential to (re)build trust, enable informed choice and respect community agency.

### Strengths & Limitations

Integrating co-production with young people and community stakeholders throughout the project, alongside service provider perspectives, enabled deeper engagement with findings and more nuanced mapping while surfacing issues at risk of being overlooked. Examining specificity of equity-oriented practices alongside the contextual realities of community care surfaced distinctions earlier mappings could not. The comprehensive search strategy and iterative integrative analysis facilitated stronger understanding of the landscape complexities. Cross-checking funding status against Charity Commission and 360Giving records provided insight into the extent of commissioning, funding cycles and surfaced the ‘independence’ grey area.

We focused on supports that identified with specific mental health and wellbeing terms, meaning some would not have met our threshold, such as spiritual counselling through places of worship. As part of our analyses we recognise the presence and purpose of these and other hidden services, though by virtue of not being findable through our scoping process we were unable to capture the scope of these services.

Given service webpages are updated on a rolling basis, provision is active with programming continually adapting, and the cyclical nature of the mapping process as services closed, went into or came out of contract, this analysis captures a snapshot in time that may not reflect current provision, nor can we infer from online information what is being actioned in practice. This study focused on two urban regions selected for their demographic and socio-political-historical contexts, while services and equity orientation likely vary in other areas (25). To meaningfully communicate findings, we clustered types of support and specialisations which inevitably masks subtleties in provision and may not reflect how services would self-identify. While we attempted to contact all service providers, many did not respond to confirm or change details, as such, our results rely on publicly available information. Limited responses likely reflect fatigue, under-resourcing, and broader frustrations with research extraction that offers little tangible benefit to services themselves.

### Conclusions & Recommendations

Community services provide vital contextually responsive care, a vast majority offering equity-oriented provision where statutory services are lagging behind. However, it’s important not to romanticise the realities, challenges exist, largely as products of systemic under-resourcing and funding power dynamics. There is no categorically ‘safe’ system, recognising safety is individual, contextual and relational. But by acknowledging the trade-offs that families are faced with, enabling and working towards cross-system collaboration, there is an opportunity to create ‘safer’ systems, improve access to suitable supports and reduce inequalities.

Policymakers should establish national guidance on budget allocation for early intervention and prevention services in children’s mental health and family wellbeing, including support that addresses drivers of need, such as experiences of violence and abuse, to interrupt cycles of escalating unmet need and pressures on costly intensive services at crises. National guidelines are needed to enable embedded partnerships between CAMHS and community organisations, addressing barriers to working relationships, reducing duplication and maximising complementarity. Data sharing agreements should be developed in partnership with community organisations rather than impose rigid generic expectations (56), with transparency in the intended purposes, particularly regarding sensitive data to minimise the risk of ‘othering’ groups in ways that perpetuate harm. Providing signposting at Charity Commission and Community Interest Company registrations to connect to service navigation infrastructure would also facilitate new services joining pre-existing databases.

Funders and commissioners need to establish longer-term funding opportunities with adequate oversight, guidance and capacity-building (56) to enable strategic planning, staff retention and training, and capture meaningful impact (44). This is particularly crucial for specialised services supporting marginalised communities, as funding priorities shift in response to political climate leaving these services operating with even greater precarity. Funding calls and commissioning contracts must be developed in partnership with community providers and those with lived experience to ensure responsiveness to community-identified gaps and needs. Ring-fencing funding for smaller grassroots services and rewarding partnerships between community organisations would also protect against larger organisations dominating funding to the detriment of hyperlocal supports engaging with targeted communities.

Across services, there is a need to move beyond acknowledging diverse needs to actively embedding anti-oppressive practices within service delivery, becoming not just provisionally responsive but organisationally oriented to empower those they serve. Critical reflection on communication practices, particularly regarding scope limitations, the nature and extent of government involvement as well as use of generic ‘keywords’ is needed; specifying practices, resources available, the criteria a child needs to meet to safely engage or the needs the service can support, would better serve inclusivity.

Several directions for future research stem from our work. Examining families’ experiences of accessing community-based and statutory services, alongside providers’ perceptions of cross-system working, would provide insight into what each system offers and misses – and contribute to the development of workable guidance. Mapping and monitoring the implementation of the NHS PCREF in CAMHS would provide insight into barriers impeding the integration of equity-oriented guidance in statutory services, which community partners may be suited to addressing. Additional evaluations of equity-oriented practices within commissioned services would extend the picture our analysis captured.

## Data Availability

All data produced in the present study are available upon reasonable request to the authors.

## Acknowledgements

We would like to express our appreciation to additional and former stakeholder advisors (in addition to those who are named authors). Their meaningful contributions and thoughtful insights shaped this project, contextualisation and co-developed recommendations.

We gratefully acknowledge all service providers who shared insights and recommendations. In particular, we extend our wholehearted gratitude to Kids on the Green, Unique Community and Migrants Organise for their invaluable reflections.

